# mRNA-1273 but not BNT162b2 induces antibodies against polyethylene glycol (PEG) contained in mRNA-based vaccine formulations

**DOI:** 10.1101/2022.04.15.22273914

**Authors:** Juan Manuel Carreño, Gagandeep Singh, Johnstone Tcheou, Komal Srivastava, Charles Gleason, Hiromi Muramatsu, Parnavi Desai, Judith A. Aberg, Rachel L. Miller, PARIS study group, Norbert Pardi, Viviana Simon, Florian Krammer

## Abstract

Two messenger RNA (mRNA)-based vaccines are widely used globally to prevent coronavirus disease 2019 (COVID-19). Both vaccine formulations contain PEGylated lipids in their composition, in the form of polyethylene glycol [PEG] 2000 dimyristoyl glycerol for mRNA-1273, and 2 [(polyethylene glycol)-2000]-N,N-ditetradecylacetamide for BNT162b2. It is known that some PEGylated drugs and products for human use that contain PEG, are capable of eliciting immune responses, leading to detectable PEG-specific antibodies in serum. In this study, we determined if any of the components of mRNA-1273 or BNT162b2 formulations elicited PEG-specific antibody responses in serum by enzyme linked immunosorbent assay (ELISA). We detected an increase in the reactivity to mRNA vaccine formulations in mRNA-1273 but not BNT162b2 vaccinees’ sera in a prime-boost dependent manner. Furthermore, we observed the same pattern of reactivity against irrelevant lipid nanoparticles from an influenza virus mRNA formulation and found that the reactivity of such antibodies correlated well with antibody levels against high and low molecular weight PEG. Using sera from participants selected based on the vaccine-associated side effects experienced after vaccination, including delayed onset, injection site or severe allergic reactions, we found no obvious association between PEG antibodies and adverse reactions. Overall, our data shows a differential induction of anti-PEG antibodies by mRNA-1273 and BNT162b2. The clinical relevance of PEG reactive antibodies induced by administration of the mRNA-1273 vaccine, and the potential interaction of these antibodies with other PEGylated drugs remains to be explored.

## Introduction

Since their introduction, vaccines currently used to prevent coronavirus disease 2019 (COVID-19) have been extensively scrutinized with regard to their safety profile. Some individuals developed adverse reactions following the vaccine administration, such as pain, itching, redness, swelling, and induration at the injection site, or general adverse reactions including cough, diarrhea, fatigue, fever, and headache (1, 2). These side effects are unpleasant but generally are not clinically serious. More severe but very rare side effects, including myocarditis and pericarditis (40 cases per million vaccinated male 12-29 year old vaccinees) have been described (3-5) and anaphylaxis (0.001 - 0.0001 % of the vaccinated population) also has been reported (2). Importantly, higher self-reported reactogenicity following administration of mRNA-1273 (from Moderna) versus BNT162b2 (from Pfizer) has been described (6). Delayed large local reactions, which are harmless but can be concerning for vaccinees, have been reported specifically to occur in mostly female vaccinees who received mRNA-1273 (7, 8). However, so far, the cause of these differences between the two mRNA-based vaccine formulations remains unclear.

It is known that some of the excipients contained in different drugs for human use can cause local or systemic reactions, resulting in the induction of immune responses towards some of these components (9). Furthermore, it has been suggested that certain components of the mRNA vaccine formulations might be involved in the development of some of the adverse reactions observed, including anaphylaxis (2, 9). Pertaining to the structure and composition of the two mRNA-based vaccine formulations, both vaccines consist of nucleoside-modified mRNA which encodes a diproline (2P)-stabilized, full-length, membrane-bound spike protein (3). Likewise, both formulations contain charged and non-charged ionizable lipids, which form the core of the lipid nanoparticles (LNPs), while 1,2-distearoyl-sn-glycero-3-phosphocholine (DSPC), cholesterol and a PEGylated lipids, give shape and stabilize the surface lipid bilayer (3) (**Table 1**). Particularly, polyethylene glycol (PEG) contained in mRNA-based formulations may be linked to some cases of anaphylaxis (10, 11). Indeed, the US Advisory Committee on Immunization Practices (ACIP) recommended in the past the exclusion of people with known severe allergic reactions against PEG and related compounds from receiving vaccine formulations containing these components (12). Both, PEG and polysorbate 80, a component of the adenovirus vectored vaccine AZD1222, as well as other compounds of similar nature, have been implicated in rare cases of hypersensitivity reactions (9, 10, 13), and PEG is known to elicit antigen-specific antibody responses in some individuals (10, 13-15).

**Table 1.** Components of mRNA-1273 and BNT162b2 formulations.

|  | mRNA-1273 | BNT162b2 |
| --- | --- | --- |
| Nucleic acid | messenger ribonucleic acid (mRNA) | messenger ribonucleic acid (mRNA) |
| Lipidic components | SM-102 | ALC 315 |
|  | polyethylene glycol [PEG] 2000<br>dimyristoyl glycerol [DMG] | 2 [(polyethylene glycol)-2000]-N,N-ditetradecylacetamide |
|  | Cholesterol | Cholesterol |
|  | 1,2-distearoyl-sn-glycero-3-phosphocholine [DSPC] | 1,2-Distearoyl-sn-glycero-3-phosphocholine [DSPC] |
| Other components | Tromethamine, tromethamine hydrochloride, acetic acid, sodium acetate trihydrate, and sucrose | Tromethamine, tromethamine hydrochloride, and sucrose or* potassium chloride, monobasic potassium phosphate, sodium chloride, dibasic sodium phosphate dihydrate, and sucrose |

\*BNT162b2 vaccine for individuals 12 years of age and older contain one of the two sets of additional ingredients

Given that both of the current mRNA-based vaccine formulations - mRNA-1273 and BNT162b2 - contain PEG in their composition, but vaccinated individuals display differential reactogenicity, we evaluated whether serum antibodies from mRNA-based vaccine recipients were able to react with homologous/heterologous mRNA vaccine formulations. Furthermore, we assessed if differential patterns of reactivity of antibodies elicited by mRNA-1273 or BNT162b2 were observed, and assessed the target components within the vaccine formulation.

## Materials and methods

### Study participants and human samples

Samples were collected as part of our ongoing institutional review board–approved, longitudinal observational studies (Study ID-20-00442: 64 participants; Study ID-16-01215: one participant). The majority of samples were selected from the PARIS (Protection Associated with Rapid Immunity to SARS-CoV-2) study, which follows healthcare workers (HCWs) of the Mount Sinai Health System. All participants signed informed consents prior to data and sample collection. Information on SARS-CoV-2 mRNA vaccine-associated side-effects were collected using a survey sent to the participants after the first and second vaccine dose. Samples were coded and analyzed in a blinded manner.

Sixty longitudinal samples from 20 PARIS participants (10 BNT162b2 vaccinees and 10 mRNA-1273 vaccinees) were selected at baseline, 18.9 days (arithmetic mean ±2.4 SD) after the first SARS-CoV-2 mRNA vaccine dose (prime) and 19.3 days (arithmetic mean ±3.9 SD) after the second SARS-CoV-2 mRNA vaccine dose (boost).

In addition, we selected longitudinal serum samples collected from participants who reported experiencing more pronounced, unusual or delayed onset vaccine-associated side effects (7 days arithmetic mean ±5.2 SD) after the first mRNA vaccine dose. The sera were collected at baseline (n=28 for BNT162b2 and n=19 for mRNA-1273 groups), 14.5 days (arithmetic mean ±4.8 SD) after the prime (n=23 for BNT162b2 and n=17 for mRNA-1273 groups) and 25.2 days (arithmetic mean ±11.6 SD) after the boost (n=26 for BNT162b2 and n=17 for mRNA-1273 groups).

The SARS-CoV-2 mRNA vaccine side effects reported from the participants selected based on their vaccine associated side effects were generally mild to moderate and self-limiting (e.g., injection site pain/swelling, fever, fatigue, etc.) with a subset of participants (N: 8, all females, three received BNT162b2 and five received mRNA-1273) reporting delayed onset, injection site rashes, redness or swelling. Of note, all five participants receive a second vaccine dose and the delayed onset skin reactions did not re-occur. Sera from a female participant who developed a severe allergic reaction after the first BNT162b2 vaccine dose requiring hospitalization were also included. This participant did not receive a second vaccine dose.

### Irrelevant mRNA-LNP production

The mRNA used as an irrelevant control was designed based on the influenza virus B/Colorado/06/2017 neuraminidase (NA) sequence. Production of the mRNA was performed as described earlier (16, 17). Briefly, the codon-optimized NA gene was synthesized (Genscript) and cloned into an mRNA production plasmid. A T7-driven *in vitro* transcription reaction (Megascript, Ambion) using linearized plasmid template was performed to generate mRNA with 101 nucleotide long poly(A) tail. Capping of the mRNA was performed in concert with transcription through addition of a trinucleotide cap1 analogue, CleanCap (TriLink) and m1Ψ-5’-triphosphate (TriLink) was incorporated into the reaction instead of UTP. Cellulose-based purification of NA mRNA was performed as described (18). The mRNA was then tested on an agarose gel before storing at -20°C.

The cellulose-purified m1Ψ-containing NA mRNA was encapsulated in LNPs using a self-assembly process as previously described wherein an ethanolic lipid mixture of ionizable cationic lipid, phosphatidylcholine, cholesterol and polyethylene glycol-lipid was rapidly mixed with an aqueous solution containing mRNA at acidic pH (19). The RNA-loaded particles were characterized and subsequently stored at - 80°C at a concentration of 1 mg/ml.

### Expression and purification of recombinant SARS-CoV-2 spike protein

Recombinant SARS-CoV-2 spike protein was produced using a mammalian cell protein expression system. Briefly, the spike (S) gene sequence (GenBank: MN908947) was cloned into a mammalian expression vector pCAGGs, as described (20, 21). Protein was expressed using the Expi293 Expression System (Thermo Fisher Scientific), according to the manufacturer’s instructions. Cell culture supernatant was collected and clarified by centrifugation at 4000 x g, filtered, and purified with Ni-nitrilotriacetic acid (NTA) agarose (QIAGEN). The purified protein was concentrated using Amicon Ultracell (Merck Millipore) centrifugation units, and the buffer was exchanged to a phosphate buffer solution (PBS, pH 7.4). Proteins were stored at -80°C until use.

### Enzyme linked immunosorbent assay (ELISA)

Antibody titers in vaccinees’ sera were determined against the recombinant trimeric spike protein of wild type SARS-CoV-2 as previously described (21, 22). Spike and BSA ELISAs were performed using phosphate-buffered saline (PBS) with 0.1% Tween-20 (PBS-T; Fisher Scientific) in washing, blocking, and diluting solutions. mRNA vaccines BNT162b2 (from Pfizer) and mRNA-1273 (from Moderna), irrelevant mRNA LNPs, multi-PEGylated bovine serum albumin (mPEG-BSA, 20 kDa, Life Diagnostics, Inc), and low molecular weight PEG (3.35kDa, Sigma) based ELISAs were performed using a modified protocol in which Tween-20 was excluded from washing/Ab solutions. Briefly, polystyrene 96-well microtiter plates (Thermo Fisher Scientific) were coated overnight with mPEG-BSA (2μg/ml), BNT162b2 or mRNA-1273 vaccine formulations (25μl/10ml), irrelevant mRNA lipid nanoparticles (0.5μg/ml) or BSA (1% solution, MP Biomedicals). For IgE and BSA controls (shown in Supplementary Figure 2), ELISA plates were coated with 2μg/ml of an IgE isotype control (Invitrogen) and BSA (1% solution, MP Biomedicals) respectively. The following day, wells were washed and blocked with 200 ul of 3% non-fat milk (AmericanBio) in PBS for 1 h at room temperature (RT). After 1 h incubation, blocking solution was removed and pre-diluted sera (in PBS 1% non-fat milk) were added at an initial dilution of 1:50 followed by 2-fold serial dilutions. After 2 h incubation, plates were washed three times with PBS and then incubated for 1 hour with anti-human IgG (Fab-specific) horse radish peroxidase (HRP) secondary antibody produced in goat (Sigma-Aldrich), or IgM-HRP (Southern Biotech) at a 1:3000 in 1% milk PBS. Specific spike/PEG-IgE antibodies were assessed by incubating with an anti-IgE HRP conjugated antibody (Invitrogen) for 1 h at a 1:2000 dilution. For the IgE control, plates were incubated with serial dilutions (2-fold) of the anti-IgE HRP conjugated antibody (Invitrogen) starting at a 1:1000 dilution for 1 h. For the BSA control, plates were incubated with serial dilutions (2-fold) of an anti-albumin (bovine serum) rabbit IgG fraction (anti-BSA, Invitrogen) starting at a 1:1000 dilution for 1 h, followed by three washes with PBS and addition of a donkey anti-rabbit IgG HRP conjugated antibody (CiteAb) at a 1:1000 dilution. Plates were washed three times with PBS and 100μl/well of *O*-phenylenediamine dihydrochloride (OPD) substrate (SigmaFast OPD; Sigma-Aldrich) were added. After 10 min incubation at RT, the reaction was stopped by addition of 50 μl of 3 M HCl solution. The optical density (OD) was measured at 490 nm using a Synergy 4 plate reader (BioTek). Data were captured in excel and are urea under the curve (AUC) values were determined using Prism 9 (GraphPad Software, San Diego, CA, USA).

### Statistical analyses

Data plotting and statistical analyses were performed using GraphPad Prism 9 (GraphPad Software, San Diego, CA, USA). Statistically significant differences between post-prime/post-boost vs baseline antibody levels were measured using a one-way ANOVA with Tukey’s multiple comparisons test. All adjusted P values of <0.05 were considered statistically significant with a confidence interval of 95%.

## Results

### mRNA-1273 vaccination induces antibodies against mRNA vaccine formulations in a prime-boost dependent manner

It has been hypothesized that the adverse reactions observed in some individuals after administration of the currently available mRNA vaccines might be caused by some of the formulation components (2, 11). To explore if vaccinees elicited antibodies against components of the vaccine formulation, we used samples from individuals who received either the mRNA-1273 or BNT162b2 mRNA vaccines. Samples were collected at baseline (n=10/group), 18.9 days (arithmetic mean ±2.4 SD) after the prime (n=10/group) or 19.3 days (arithmetic mean ±3.9 SD) after the boost (n=10/group). Initially, to confirm the induction of antibodies after the vaccine administration, we measured the IgG titers against a recombinant version of the spike protein via ELISA. We detected a prime-boost dependent induction of anti-spike antibodies after vaccination, with levels oscillating around 10^3^ area under the curve (AUC) units after prime with mRNA-1273 or BNT162b2, and 10^4^ after the boost administration (Figs. **1A** and **1B**). Then, we coated ELISA plates with a standard amount of each of the vaccine formulations resuspended in PBS and assessed binding of IgG antibodies. We found that sera collected after vaccination with mRNA-1273 had increasing reactivity against both the BNT162b2 (Fig. **1C**) and mRNA-1273 (Fig. **1D**) formulations, and that the increased reactivity was vaccination-dependent, with a moderate increase after the prime administration and higher levels induced after the boost. Interestingly, this increase in reactivity was not evident in sera from individuals receiving the BNT162b2 vaccine, either against the BNT162b2 or mRNA-1273 formulations (Figs. **1E** and **1F**). Overall, these data suggest that mRNA-1273 but not BNT162b2 vaccination induces antibodies against some component(s) of the vaccine formulations.

**Figure 1.**
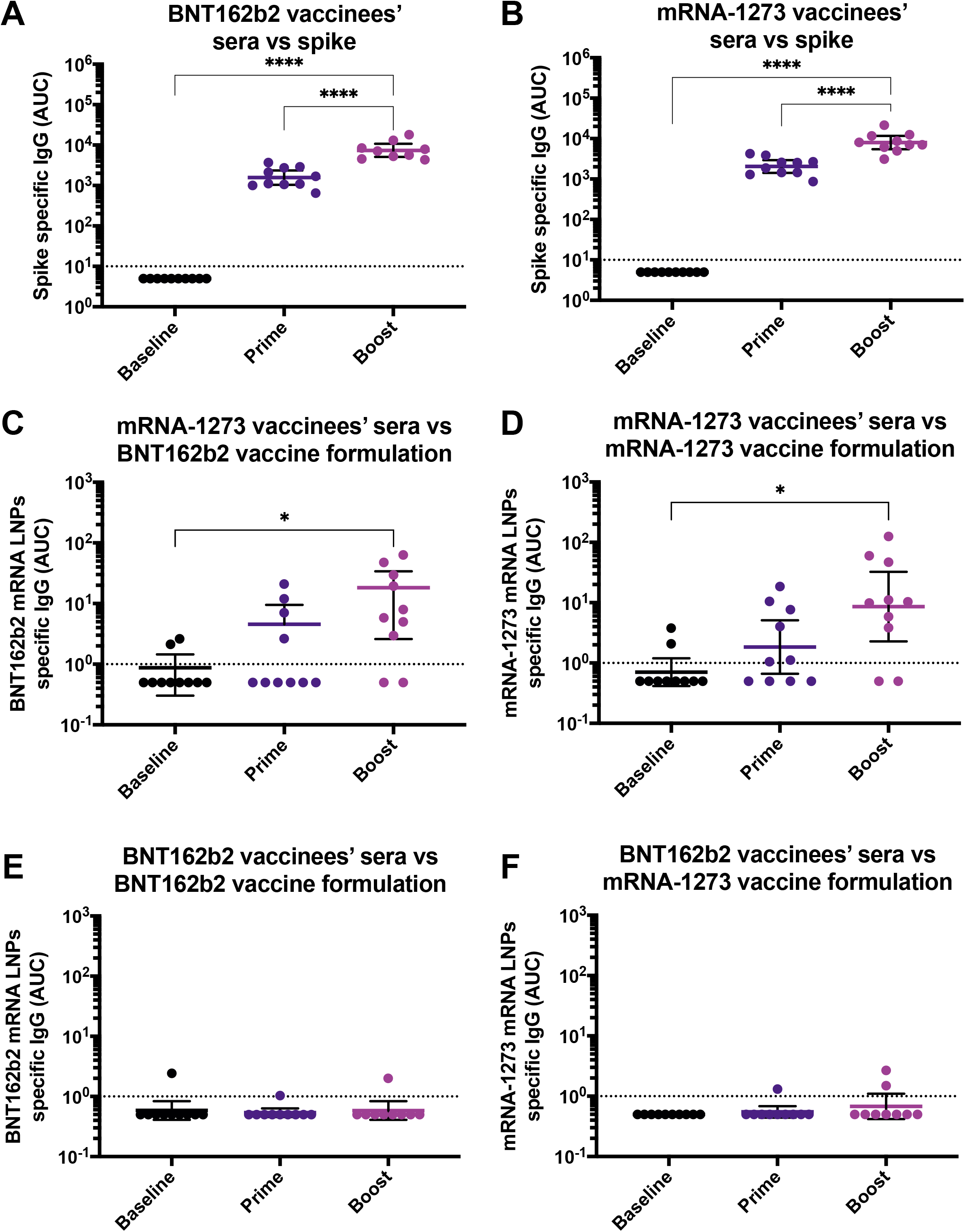
Antibodies against SARS-CoV-2 spike and mRNA-based vaccine formulations in vaccinees’ sera. Sera from BNT162b2 (left column) or mRNA-1273 (right column) vaccinees collected at baseline (n=10 for BNT162b2 and n=10 for mRNA-1273 groups), 18.9 days (arithmetic mean ±2.4 SD) after the prime (n=10 for BNT162b2 and n=10 for mRNA-1273 groups) and 19.3 days (arithmetic mean ±3.9 SD) after the boost (n=10 for BNT162b2 and n=10 for mRNA-1273 groups), were tested for IgG antibodies against SARS-CoV-2 full-length spike (**A** and **B**), BNT162b2 mRNA LNPs (**C** and **E**), and mRNA-1273 mRNA LNPs (**D** and **F**) by ELISA. Dotted line represents the limit of detection (LOD) of the assay. Statistically significant differences between post-prime/post-boost vs baseline antibody levels are shown. One-way ANOVA with Tukey’s multiple comparisons test. *, P < 0.05; **, P < 0.01; ***, P < 0.001; ****, P < 0.0001.

### Antibodies reactive towards the vaccine formulation are directed against the lipid nanoparticles and react with polyethylene glycol (PEG)

Lipid nanoparticles contained in the currently used mRNA vaccine formulations, as well as other drugs and other cosmetic and health products for human use, contain PEG and have the potential to elicit immune responses against it (10, 13-15). To dissect the target within the vaccine formulation to which mRNA-1273-induced antibodies bind, we coated ELISA plates with LNPs carrying a SARS-CoV-2 unrelated, irrelevant mRNA (encoding influenza virus neuraminidase). Similar to the reactivity overserved against the BNT162b2 and mRNA-1273 formulations, we detected an increase in the reactivity against the irrelevant mRNA-LNPs in individuals receiving the mRNA-1273 vaccine (Fig. **2B**), but not in those ones vaccinated with BNT162b2 (Fig. **2A**), suggesting the reactivity is independent of the sequence of the mRNA contained in the formulation.

**Figure 2.**
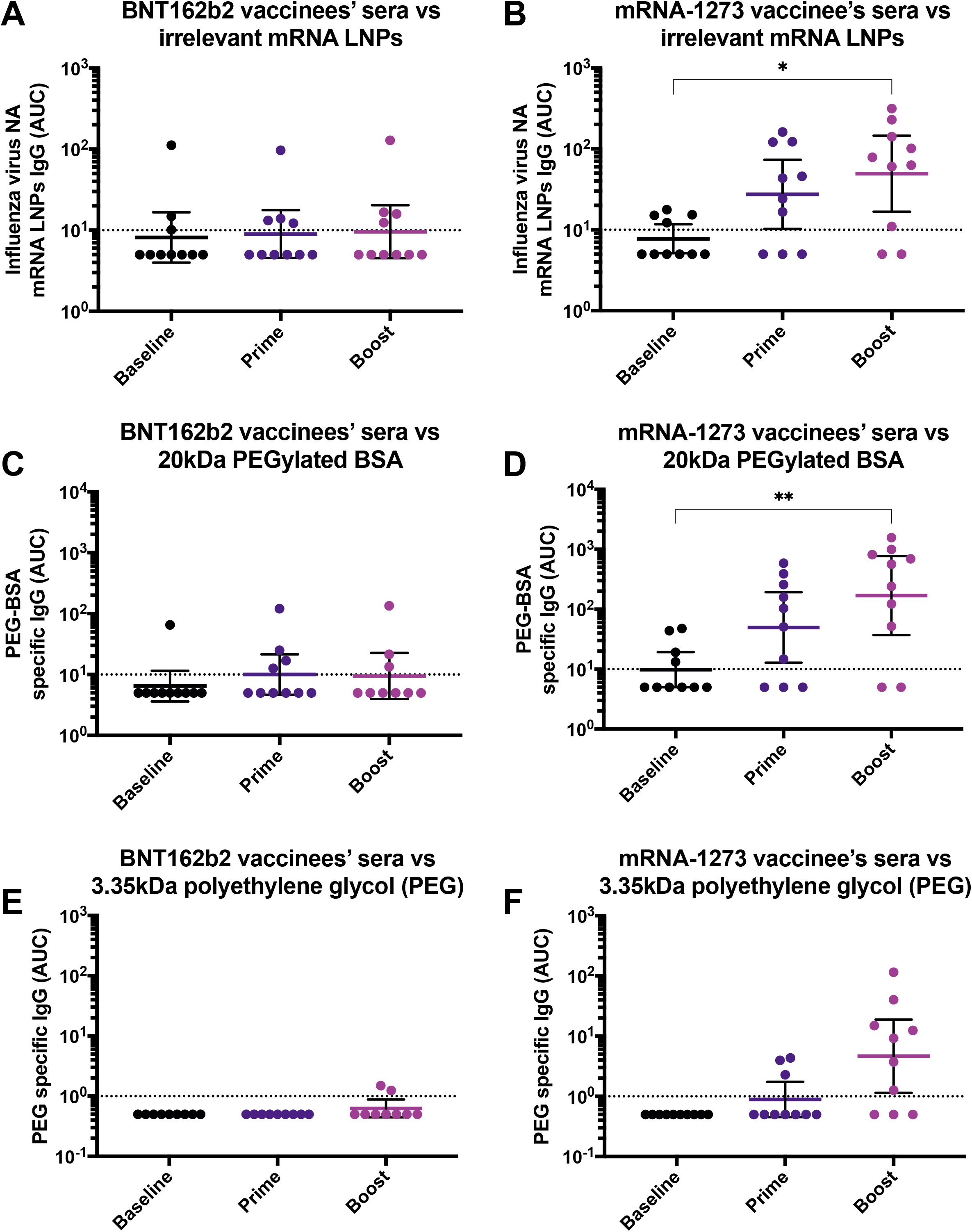
Antibodies against irrelevant lipid nanoparticles (LNPs) and polyethylene glycol (PEG) in vaccinees’ sera. Sera from BNT162b2 (left column) or mRNA-1273 (right column) vaccinees collected at baseline (n=10 for BNT162b2 and n=10 for mRNA-1273 groups), 18.9 days (arithmetic mean ±2.4 SD) after the prime (n=10 for BNT162b2 and n=10 for mRNA-1273 groups) and 19.3 days (arithmetic mean ±3.9 SD) after the boost (n=10 for BNT162b2 and n=10 for mRNA-1273 groups), were tested for IgG antibodies against irrelevant LNPs (**A** and **B**), PEGylated BSA 20kDa (**C** and **D**), 3.35kDa PEG (**E** and **F**) by ELISA. Dotted line represents the limit of detection (LOD) of the assay. Statistically significant differences between post-prime/post-boost vs baseline antibody levels are shown. One-way ANOVA with Tukey’s multiple comparisons test. *, P < 0.05; **, P < 0.01; ***, P < 0.001; ****, P < 0.0001.

Next, we assessed whether vaccination induced antibodies reacted to PEG. We measured the binding of sera from mRNA-1273 and BNT162b2 vaccinated individuals to a PEGylated form of BSA (PEG-BSA) containing high molecular weight PEG (20kDa). Again, we found that individuals vaccinated with mRNA-1273, had an increase in antibodies against PEG-BSA in a vaccination-dependent manner (Fig. **2D**), whereas no significant increase of antibody titers in individuals receiving the BNT162b2 vaccine was observed (Fig. **2C**). As an alternative experimental approach, we coated high binding polystyrene plates directly with a low molecular weight PEG (3.35 kDa) molecule and performed similar ELISAs. Although this method seemed to be less sensitive than the PEG-BSA ELISA, similarly we observed that individuals vaccinated with mRNA-1273, had increased reactivity to PEG, particularly after the boost (Fig. **2E**), but there was no increase in antibody titers following BNT162b2 administration (Fig. **2F**). Overall, these results suggest that the antibodies induced towards the vaccine formulation components may be directed against PEG, which is present in the vaccine formulation.

### PEG as the target of mRNA-1273-induced antibodies

To assess in a systematic manner if the formulation-reactive antibodies induced in mRNA-1273 vaccine recipients were directed against PEG, we performed correlation analyses using the AUC values obtained in the different ELISAs performed. We found that the mRNA-1273 induced antibodies detected against the BNT162b2 (Fig. **3A**) or mRNA-1273 (Fig. **3B**) vaccine formulations not only correlated well with PEG as measured by using PEGylated BSA, but a correlation was observed between the PEGylated-BSA AUC values and the irrelevant mRNA-LNPs (Fig. **3C**), as well as with the low molecular weight PEG (3.35kDa) AUCs (Fig. **3D**). Moreover, the independent correlation of PEGylated-BSA AUC values vs the BNT162b2 or mRNA-1273 formulation, the irrelevant mRNA-LNPs, or the low molecular weight PEG AUCs, increased in a prime-boost dependent manner, with the lowest correlation observed at baseline, and increasing correlations after the prime, followed by high correlations after the boost (Supplementary Fig. **1**). The absolute values of the geometric mean AUCs for every ELISA and the fold induction after the prime or the boost are shown in **Table 2**. Overall, these analyses support that the antibodies detected in mRNA-1273 vaccine recipients, which react towards the BNT162b2 or mRNA-1273 vaccine formulations, are directed towards the PEG component of the formulations.

**Table 2.** Geometric means of AUC and AUC fold change for each ELISA antigen.

|  |  | Geometric mean AUCs |  | Geometric mean AUC fold change |  |
| --- | --- | --- | --- | --- | --- |
|  | Timepoint/<br>vaccine | BNT162b2<br>G.M. AUC | mRNA-1273<br>G.M. AUC | BNT162b2<br>(G.M.F.I) | mRNA-1273<br>(G.M.F.I) |
| **Spike** | Prime | 1569.32 | 2043.64 | 313.86 | 408.73 |
|  | Boost | 7405.92 | 7974.32 | 1481.18 | 1594.86 |
| **BNT162b2 LNP** | Prime | 0.54 | 1.54 | 0.92 | 2.25 |
|  | Boost | 0.58 | 7.04 | 1.07 | 4.28 |
| **mRNA-1273 LNP** | Prime | 0.55 | 1.84 | 1.1 | 2.6 |
|  | Boost | 0.66 | 8.62 | 1.32 | 12.2 |
| **Nonspecific LNP** | Prime | 8.98 | 27.44 | 1.1 | 3.54 |
|  | Boost | 9.6 | 49.39 | 1.07 | 2.12 |
| **PEGylated BSA** | Prime | 10.01 | 49.64 | 1.26 | 5.06 |
|  | Boost | 8.87 | 169.14 | 1.11 | 17.25 |
| **Low molecular weight PEG** | Prime | 0.5 | 0.89 | 1 | 1.78 |
|  | Boost | 0.61 | 1.78 | 1.22 | 9.29 |
| **BSA** | Prime | 5.43 | 5.38 | 0.92 | 0.96 |
|  | Boost | 5.78 | 8.06 | 0.98 | 1.44 |

Geometric mean (G.M.) of all the area under the curve (AUC) values measured after the prime or the boost are shown in the left panels. Geometric mean of fold induction (G.M.F.I) values, expressed as the ratio between prime or boost AUCs and the baseline AUC for each of the participants is shown in the right panels. Prime: First SARS-CoV-2 vaccine dose; Boost: Second SARS-CoV-2 vaccine dose

**Figure 3.**
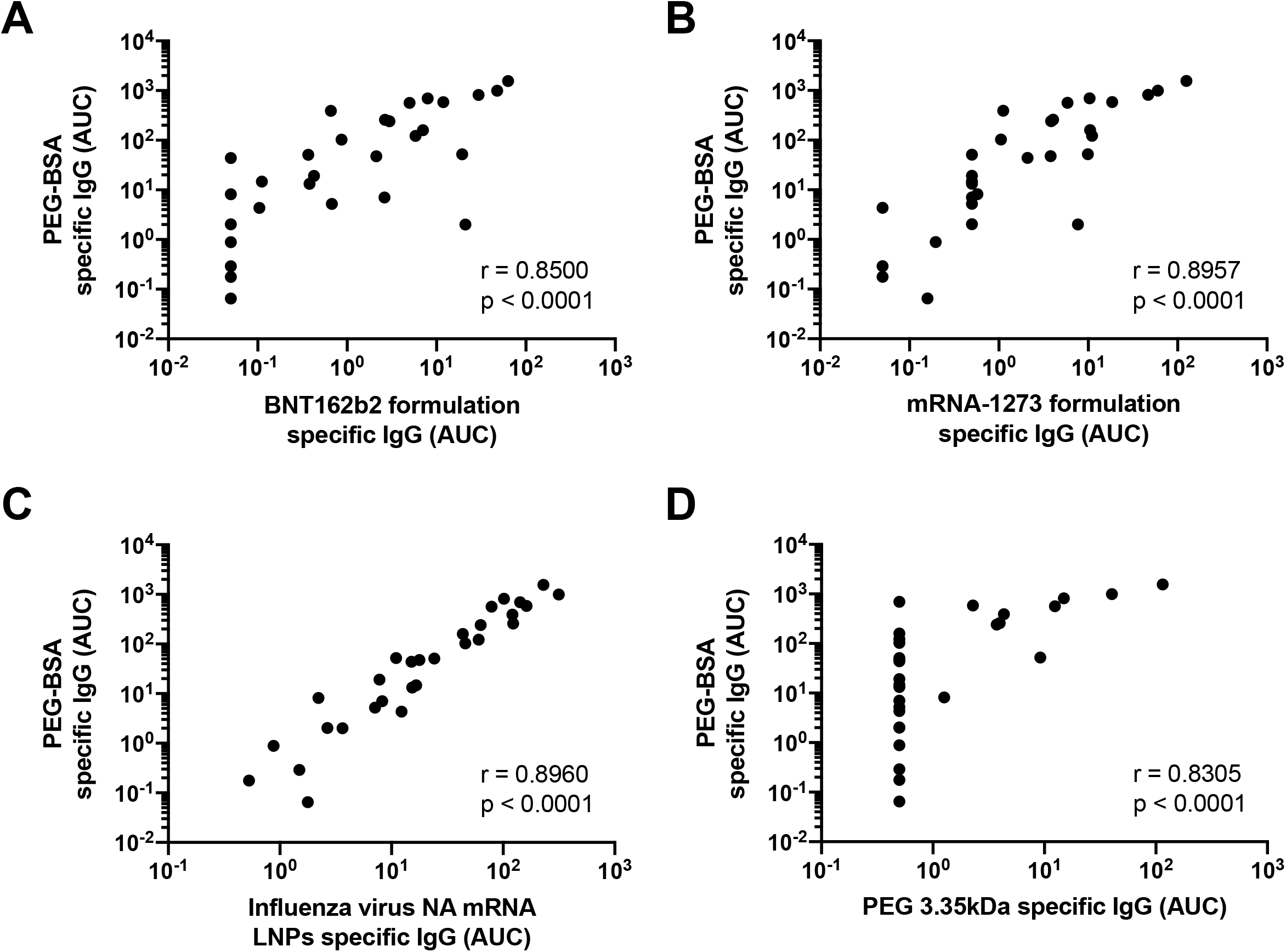
Correlation of polyethylene glycol (PEG) reactive antibodies among different assays. Area under the curve (AUC) values obtained from the 20kDa PEGylated-BSA ELISA using mRNA-1273 sera were analyzed for correlation with BNT162b2 formulation specific IgG (**A**), mRNA-1273 formulation specific IgG (**B**), irrelevant LNPs IgG (**C**), and PEG 3.35KDa specific IgG (**D**). Pearson correlation was used. Statistically significant differences between post-prime/post-boost vs baseline antibody levels are shown. One-way ANOVA with Tukey’s multiple comparisons test. *, P < 0.05; **, P < 0.01; ***, P < 0.001; ****, P < 0.0001.

To explore if vaccinees displayed other classes of anti-PEG antibodies, we measured the reactivity of IgM using the PEGylated BSA based ELISA. Similar to the IgG pattern previously observed, we detected PEG-specific IgM - although at low levels - in the mRNA-1273 recipients in a vaccination dependent manner (Fig. **4B**), however no induction of IgM in the BNT162b2 vaccinees (Fig. **4A**). Moreover, we assessed whether individuals could potentially induce IgE antibodies directed to PEG in response to vaccination, however levels of PEG-specific IgE were undetectable in all the participants, irrespective of the vaccine type received (Figs. **4C** and **4D**). As a control for IgE detection, we used plates coated with house dust mite (HDM) antigens, which allowed detecting IgE in some of the participants (Supplementary Fig. **2**). Moreover, as a control to ensure that the anti-PEG antibodies detected through this work were directed specifically against PEG, and exclude the possibility that the BSA contained in the PEG-BSA reagent was as a target of the reactivity detected, we performed ELISAs in plates pre-coated with 1% BSA (Figs. **4E** and **4F**). We detected residual reactivity in three individuals, although at levels very close to the limit of detection. These residual antibodies were, however, detected in both the BNT162b2 and mRNA-1273 groups, and were irrespective of the vaccination time point. Overall, these results indicate that mRNA-1273 administration leads to the induction of specific IgG and IgM, but not IgE antibodies against PEG.

**Figure 4.**
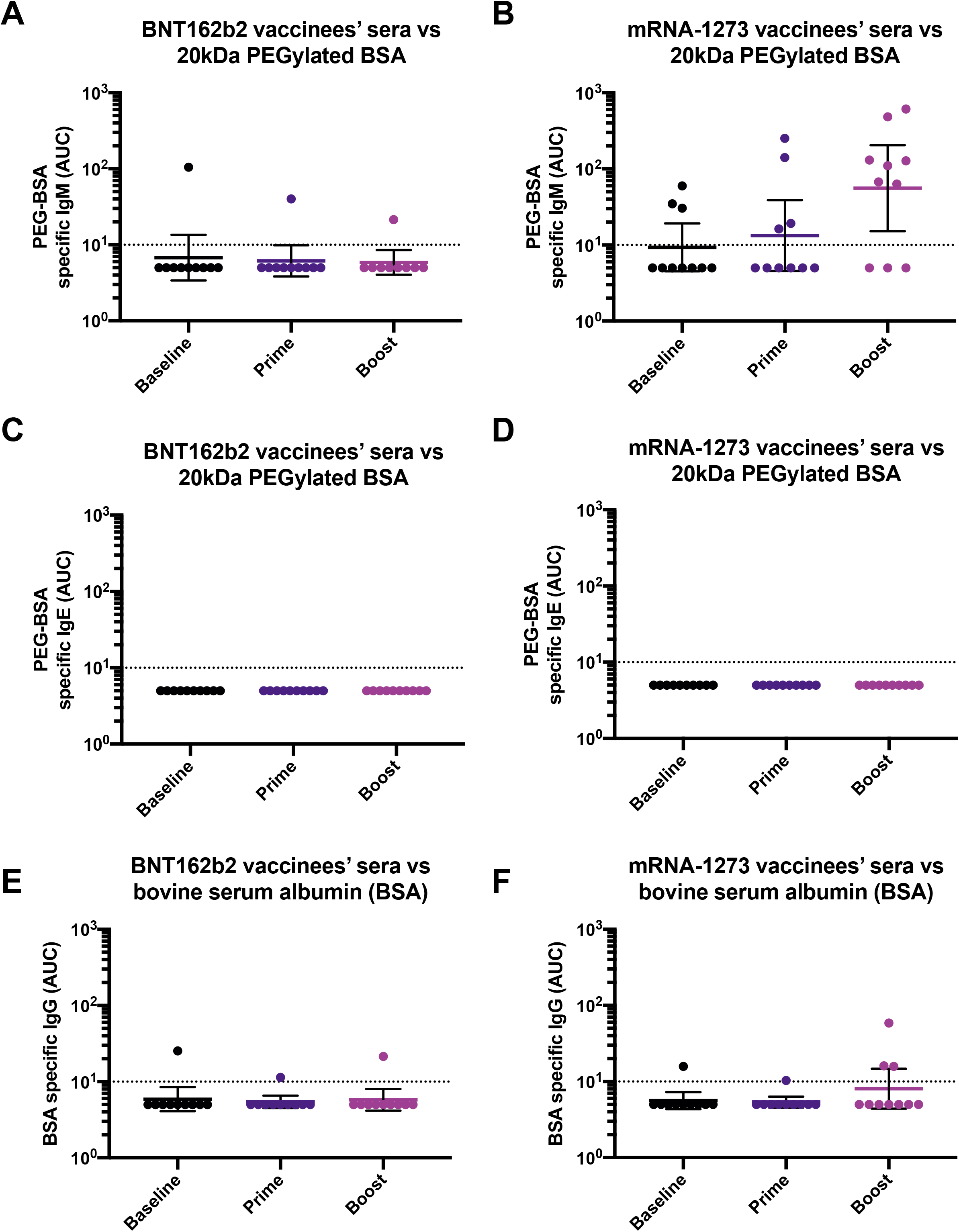
Levels of IgM and IgE antibodies against PEG and specificity of anti-PEG ELISA. Sera from BNT162b2 (left column) or mRNA-1273 (right column) vaccinees collected at baseline (n=10 for BNT162b2 and n=10 for mRNA-1273 groups), 18.9 days (arithmetic mean ±2.4 SD) after the prime (n=10 for BNT162b2 and n=10 for mRNA-1273 groups) and 19.3 days (arithmetic mean ±3.9 SD) after the boost (n=10 for BNT162b2 and n=10 for mRNA-1273 groups), were tested for IgM (**A** and **B**) or IgE (**C** and **D**) antibodies against PEGylated BSA 20kDa or against bovine serum albumin (BSA, **E** and **F**). Statistically significant differences between post-prime/post-boost vs baseline antibody levels are shown. One-way ANOVA with Tukey’s multiple comparisons test. *, P < 0.05; **, P < 0.01; ***, P < 0.001; ****, P < 0.0001.

### Anti-PEG antibodies in a subgroup of participants with reported vaccine associated side effects including delayed onset, injection site rash/erythema or severe allergic reaction

A small proportion of individuals have experienced delayed large local reactions after receiving the mRNA-1273 vaccine (7, 8). To assess whether individuals who experienced delayed onset reactions or other types of adverse reactions following vaccination, mounted differential anti-PEG antibodies at baseline or after vaccination, we used samples from a selection of study participants that reported vaccine-associated side effects such as delayed onset reactions including injection site rashes (N=8) or severe allergic reaction (N=1). Overall, although levels of anti-PEG antibodies were slightly higher at baseline, we did not find a significant association between baseline anti-PEG titers and antibody induction after vaccination with mRNA-1273 or BNT162b2. However, individuals receiving the mRNA-1273 formulation (Fig. **5B**), but not the ones receiving the BNT162b2 vaccine (Fig. **5A**), induced significantly higher levels of anti-PEG antibodies in a vaccination-dependent manner, similar to the findings described above. In summary, these findings indicate that although there is an increase in the anti-PEG antibodies in the mRNA-1273 vaccinees, there was no obvious association between PEG antibodies and adverse reactions. Pre-existing anti-PEG levels are not associated with a more robust PEG antibody induction following vaccination.

**Figure 5.**
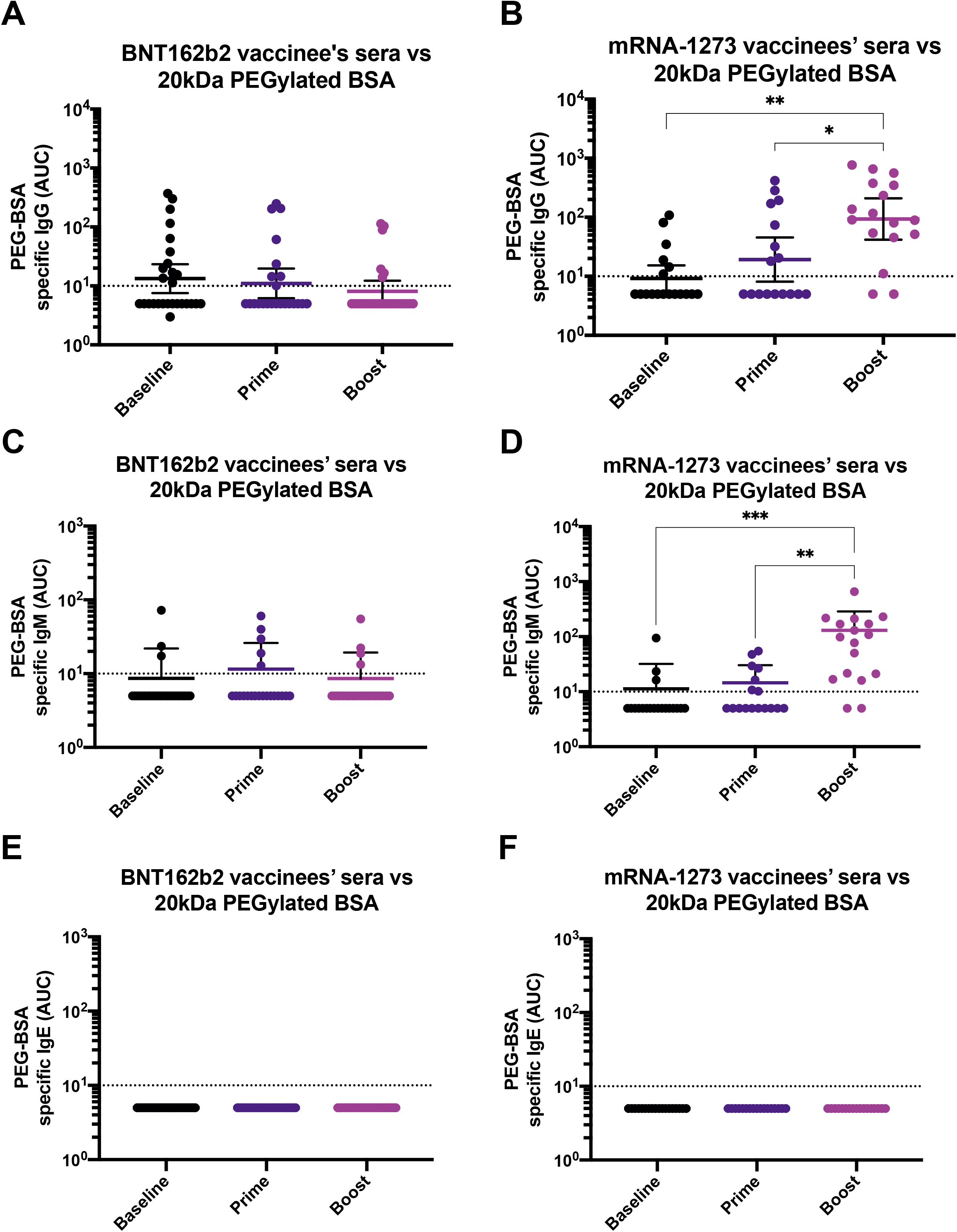
Polyethylene glycol (PEG) antibodies in participants selected based on their vaccine associated side effects including delayed onset injection site reactions and severe allergic reaction. Sera from BNT162b2 (**A**) or mRNA-1273 (**B**) vaccinees was collected at baseline (n=28 for **A and C**, and n=19 for **B and D**), 14.5 days (arithmetic mean ±4.8 SD) after the prime (n=23 for **A and C**, and n=17 for **B and D**) or 25.2 days (arithmetic mean ±11.6 SD) after the boost (n=26 for **A and C** and n=17 for **B and D**), and tested for IgG (**A** and **B**), IgM (**C** and **D**) or IgE (**E** and **F**) antibodies against 20kDa PEGylated BSA by ELISA. Dotted line represents the limit of detection (LoD) of the assay. Statistically significant differences between post-prime/post-boost vs baseline antibody levels are shown. One-way ANOVA with Tukey’s multiple comparisons test. *, P < 0.05; **, P < 0.01; ***, P < 0.001; ****, P < 0.0001.

## Discussion

Some of the current vaccines to prevent COVID-19 have unique properties as compared to any other licensed vaccines in history. They are based on mRNA encapsulated in lipid nanoparticles and they do not contain the target antigen which is coded by the mRNA and produced in the vaccinee’s cells. The LNPs contain lipidic components of diverse nature including PEGylated lipids. Polyethylene glycol (PEG) is found in different drugs, cosmetics and health products for human use (10, 23). Administration of animals with PEGylated proteins of different nature can induce PEG-specific antibodies (24). Likewise, humans are able to induce anti-PEG antibodies following administration of certain PEGylated drugs (14, 25-27). Moreover, pre-existing anti-PEG antibodies are present in some individuals and can interfere with activities of PEGylated drugs (15, 28).

Here, we found that a proportion of study participants receiving SARS-CoV-2 mRNA vaccines mount anti-PEG antibodies to variable levels. Not every study participant receiving the mRNA-1273 vaccine displayed high levels of anti-PEG antibodies post-vaccination, and not every individual experiencing delayed onset side effects such as injection site reactions post-vaccination had pre-existing anti-PEG levels. This suggests that although PEG contained in the mRNA-1273 vaccine formulation is recognized by the immune system and antibodies are induced against this molecule in some of the vaccinees, such high levels of antibodies – either at baseline or induced by vaccination – do not directly correlate with the emergence of delayed large local reactions. Although we cannot establish an association between the levels of PEG-reactive antibodies and the higher reactogenicity observed in mRNA-1273 vaccinees, our findings suggest that perhaps anti-formulation immune responses are contributing to the higher reactogenicity sometimes observed with mRNA-1273 compared to BNT162b2.

The pre-existing antibody levels against PEG could be due to previous exposures to PEGylated drugs (14, 25-27) or PEG-containing products (23). Importantly, we assessed the presence of spike- or PEG-specific IgE antibodies and we did not find detectable levels of IgE antibodies, including in the one participant who experienced a severe allergic reaction in response to vaccination. Our results are in line with previous findings detecting PEG-specific IgG following vaccination, but a lack of IgE (29). Allergy skin testing to PEG also was negative (unpublished data). Immediate allergic reactions following vaccination, such anaphylaxis, are likely to be mediated by IgE-independent mechanisms of diverse nature (29), and the relevance of PEG-specific IgG induced by vaccination remains to be investigated. Interestingly, via a genome-wide association study, an immunoglobulin heavy chain (IGH) locus has been associated with the anti-PEG IgM response (30). Although such association was not present for IgG, IGH polymorphisms associated with switched anti-PEG IgG subsets require further exploration.

Differential immunogenicity of PEG molecules containing a methoxy (mPEG), hydroxy (H)-PEG) or t-butoxy (t-BuO-PEG) distal terminal groups has been described, with mPEG being more prone to induce specific responses against this terminal group (31). Notwithstanding, the majority PEG-specific monoclonal antibodies (mAbs) recognize the backbone of the molecule, meaning the repeated ethylene oxide subunits (28, 32-34). Although the currently used mRNA formulations incorporate different forms of PEG, both the polyethylene glycol [PEG] 2000 dimyristoyl glycerol contained in the mRNA-1273 formulation, and the 2 [(polyethylene glycol)-2000]-N,N-ditetradecylacetamide contained in the BNT162b2 formulation, bear a methoxy terminal group. Hence, it is unlikely that the differential patterns of anti-PEG antibodies detected in mRNA-1273 vs BNT162b2 vaccinee’s sera, are due to PEG structural differences in the formulations, but rather, this might be the result of the higher dose of mRNA given to mRNA-1273 vaccine recipients -100μg for mRNA-1273 vs 30μg for BNT162b2, the result of the higher PEGylated lipid dose in mRNA-1273 or the way PEG is presented by the carrier lipid (35). Moreover, serum from mRNA-1273 vaccine recipients was able to recognize components of both formulations in a prime-boost dependent manner. It remains to be explored whether the anti-PEG antibodies induced following vaccination are directed towards the backbone of the PEG molecule, or the methoxy group present in PEG from both formulations. Overall, our study reports the induction of PEG antibodies following administration of one of the currently used mRNA-based vaccine formulation. The clinical relevance of PEG-reactive antibodies induced by mRNA-1273 administration and the potential interaction of these antibodies with other PEGylated drugs remain to be explored.

## Data Availability

All data produced in the present study are available upon reasonable request to the authors

## Acknowledgments

We thank the study participants for their generosity and willingness to participate in longitudinal COVID-19 research studies. None of this work would be possible without their contributions. This work is part of the PARIS/SPARTA studies funded by the NIAID Collaborative Influenza Vaccine Innovation Centers (CIVIC) contract 75N93019C00051. In addition, this work was also partially funded by NIAID U19AI168631-01 and the Centers of Excellence for Influenza Research and Surveillance (CEIRS, contract # HHSN272201400008C), the JPB Foundation, the Open Philanthropy Project (research grant 2020-215611 (5384) and by anonymous donors. Finally, this effort was also supported by the Serological Sciences Network (SeroNet) in part with Federal funds from the National Cancer Institute, National Institutes of Health, under Contract No. 75N91019D00024, Task Order No. 75N91021F00001. The content of this publication does not necessarily reflect the views or policies of the Department of Health and Human Services, nor does mention of trade names, commercial products or organizations imply endorsement by the U.S. Government. We thank Acuitas Therapeutics for LNP-encapsulating the NA mRNA used in this study.

## Author contributions

F.K., V.S. and J.M.C. conceptualized study; CG, KS and the PARIS study group enrolled participants, collected data, evaluated surveys and provided biospecimen and metadata, G.S., J.M.C, C.G., H.M., J.T., P.D. and the PARIS study group performed experiments; J.M.C., G.S., and J.T. analyzed data; J.M.C., V.S., and F.K. administered the project; F.K., V.S., and N.P. provided resources; J.M.C. wrote original draft. All authors reviewed, edited and approved the final version of the manuscript, and have had access to the raw data.

## Figure Legends

**Supplementary Figure 1. Correlation of polyethylene glycol (PEG) reactive antibodies among different assays (by vaccination time point)**. Area under the curve (AUC) values obtained from the 20kDa PEGylated-BSA ELISA using mRNA-1273 sera were analyzed for correlation with BNT162b2 formulation specific IgG, mRNA-1273 formulation specific IgG, irrelevant LNPs IgG, and 3.35KDa PEG specific IgG, at baseline (left column), after the first vaccine dose administration (central column) or after the boost (right column). Pearson correlation was used.

**Supplementary Figure 2. Controls used for measurement of IgE antibodies and for BSA ELISAs**. The positive control used for measurement of antigen-specific IgE is shown in **A**. The positive control used for measurement of BSA specific IgG is shown in **B**.

