## Supplementary figures and images for "mRNA-1273 but not BNT162b2 induces antibodies against polyethylene glycol (PEG) contained in mRNA-based vaccine formulations"

### Supplementary figures

Supplementary figure 1

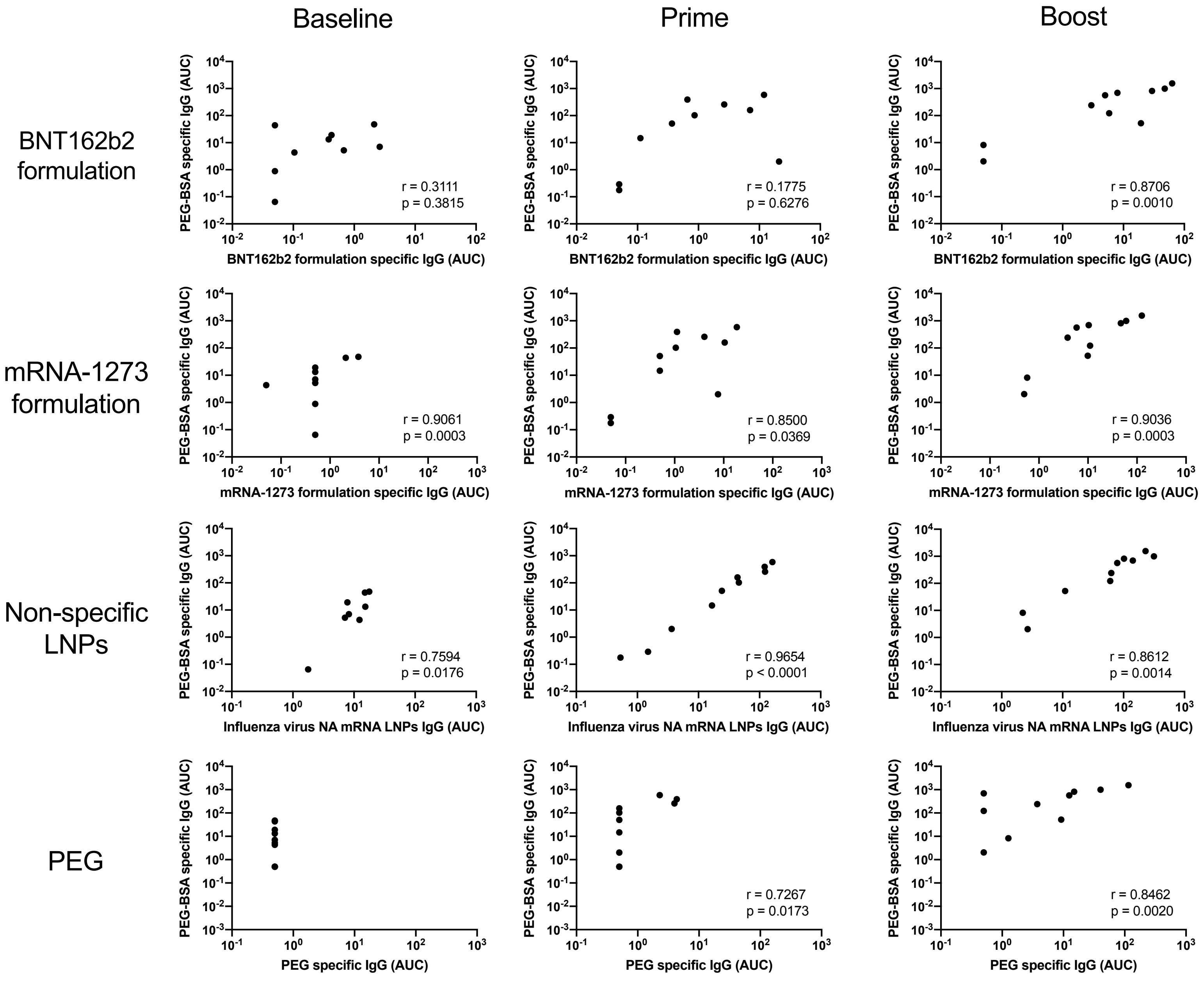

Supplementary figure 2

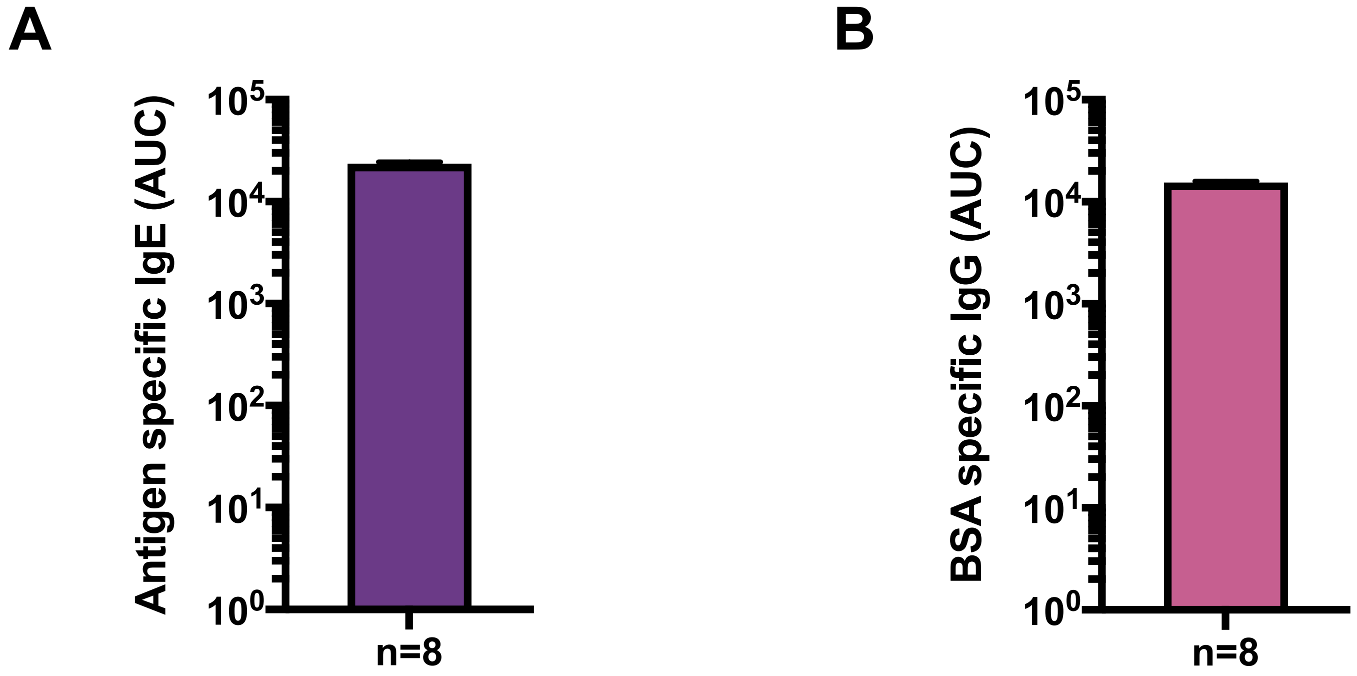
